# The PRIME-NL study: evaluating a complex healthcare intervention for people with Parkinson’s disease in a dynamic environment

**DOI:** 10.1101/2024.03.11.24304097

**Authors:** Bart R. Maas, Robin van den Bergh, Sanne W. van den Berg, Eveline Hulstein, Niek Stadhouders, Patrick P.T. Jeurissen, Nienke M. de Vries, Bastiaan R. Bloem, Marten Munneke, Yoav Ben-Shlomo, Sirwan K.L. Darweesh

## Abstract

**Background:** An innovative, integrative care model for people with Parkinson (PRIME Parkinson) has gradually been implemented in a selected region of the Netherlands since 2021. A prospective evaluation of this model (PRIME-NL study) was initiated in parallel, spanning the year prior to implementation (baseline) and the implementation period. Following publication of the original study protocol, the COVID-19 crisis delayed implementation of the full PRIME Parkinson care model by two years and hampered the recruitment of study participants.

**Objective:** To describe which methodological adjustments were made to the study protocol because of these developments.

**Methods:** We compare various outcomes between a region where PRIME Parkinson care was implemented (innovation region) versus the rest of the Netherlands (usual care region). We use healthcare claims data of virtually all people with Parkinson in the Netherlands and annual questionnaires in a representative subsample of 984 people with Parkinson, 566 caregivers and 192 healthcare professionals. Four major methodological adjustments had to be made since publication of the original protocol. First, we extended the evaluation period by two years. Second, we incorporated annual process measures of the stage of implementation of the new care model. Third, we introduced a real-time iterative feedback loop of interim results to relevant stakeholders. Fourth, we updated the statistical analysis plan.

**Discussion:** This manuscript provides transparency in how the design and analyses of the evaluation study had to be adapted to control for external influences in a dynamic environment, including eruption of the COVID-19 crisis. Our solutions could serve as a template for evaluating other complex healthcare interventions in a dynamic environment.

## 1. Introduction

Parkinson’s disease (PD) is the second most common neurodegenerative condition, and its prevalence is growing rapidly (1). The needs of people with Parkinson (PwP), including PD and atypical parkinsonism, are not met optimally by current care models. Based on the needs of PwP, their relatives and healthcare professionals, we proposed a new care model termed PRIME (Proactive and Integrated Management and Empowerment in Parkinson’s Disease) (2). The model aims to improve (I) the health of PwP, (II) the experienced quality of care by PwP and their informal caregivers and (III) the work-life balance for healthcare professionals, (IV) without raising the total costs of care. These aims are collectively referred to as the quadruple aim (3).

The new care model introduced various healthcare innovations in a stepwise fashion, which are being implemented in the South-East region of the Netherlands from 2021 onwards. The PRIME-NL study compares various outcomes between the region where this care model was implemented (innovation region) and the rest of the Netherlands (usual care region). The data sources include healthcare claims data of virtually all PwP in the Netherlands plus annual questionnaires obtained in a representative subsample of 982 PwP, 566 caregivers and 192 healthcare professionals. The rationale and design of that prospective observational evaluation was published earlier (4).

However, following the publication of the original study protocol, we faced unforeseen challenges. In the first two years since this project started, the coronavirus disease 2019 (COVID-19) outbreak emerged, which had an enormous impact on global healthcare. For example, waiting times for diagnostics and elective care increased. The COVID-19 pandemic impacted our project in various ways. First, it delayed the implementation of the new care model, because of attrition of personnel and because meetings as well as in-person pilot assessments had to be cancelled. Second, it delayed the recruitment for the questionnaire-based study, which resulted in a smaller than planned sample size for the questionnaire study. In addition, we have gained a lot of experience during implementation and evaluation period, which also resulted in additions to the study protocol.

The purpose of this paper is to outline the methodological adjustments that had to be made since the publication of the original protocol. We aim to be transparent about such modifications to enhance scientific integrity and validity. We hope that this will inspire the design of other healthcare evaluations by providing insights into real-life challenges in an observational healthcare evaluation that is being implemented and evaluated in a highly dynamic environment.

## 2. Methods

We will illustrate how the PRIME Parkinson care model is operationalized, explain the changes to the original study protocol and describe the prespecified statistical analyses in detail.

### 2.1 PRIME Parkinson care

PRIME Parkinson care is operationalized as a home, spoke and hub model (*Supplementary Material A*). This model comprises the following core elements: (a) specialized Parkinson’s nurses who operate across the patients’ total care network and play an important role in coordination and integration of care; (b) regional teams which include specialist neurologists and a network of specially trained allied health professionals (ParkinsonNet); (c) an expertise center that supports Parkinson’s nurses and regional teams; and (d) self-management by well-informed patients. This complete model is supported by various technology products and a range of centralized services, such as reliable information services, to support PwP and professionals.

The elements and services of this home, spoke and hub model are implemented in the South-East region of the Netherlands. In previous publications, we termed this region the PRIME region (4, 5). However, from now on we will call this the innovation region, because of possible confusion with the name of the evaluation study. The new care model was implemented in a stepwise fashion, starting from 2021. In *Figure 1*, we visually illustrate the timeline of implementation of all innovations of the new care model. We developed the healthcare innovations from 2019 onwards in an iterative process, adhering to principles of design thinking (6). Adopting a design thinking approach, enables PwP, caregivers and healthcare professionals to give input throughout the entire development process, which has led to changes in the intervention during the project phase.

**Fig 1.**
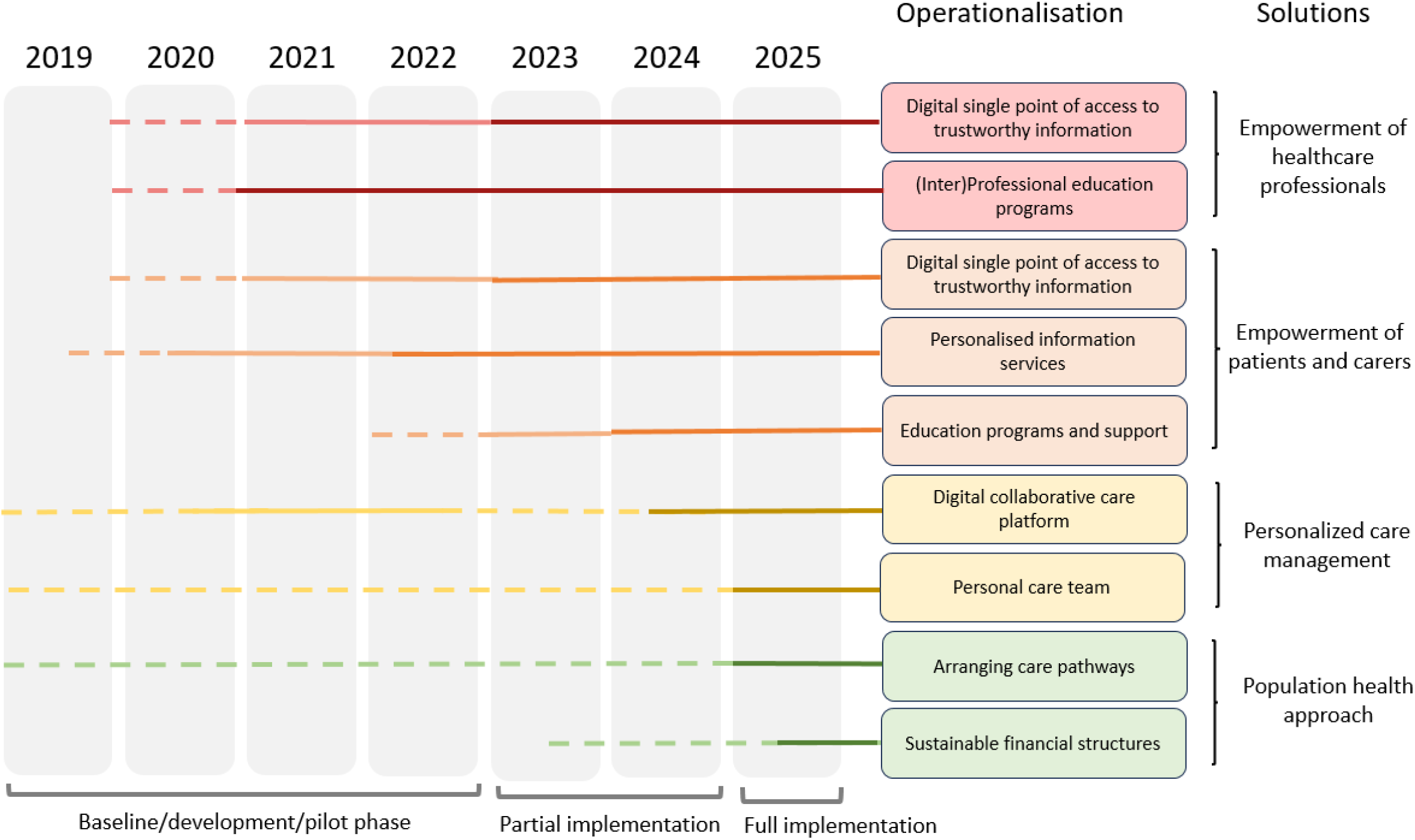
Timeline of development and implementation of the PRIME Parkinson care model. Dotted lines indicate development period, light-colored lines indicate pilot implementation in a subgroup and dark-colored lines indicate full implementation of the innovation in the innovation region. Full implementation indicates that the innovation is available to all people with Parkinson’s disease in the PRIME care region. Innovations are linked to the solutions of the logic model (3).

### 2.2 Changes to the initial protocol

We made several changes to the initial protocol. Four major methodological adjustments were made since publication of the original protocol. First, the evaluation period has been extended by two years. Second, we incorporated annual process measures of the stage of implementation of the new care model. Third, a real-time iterative feedback loop of interim results to stakeholders has been effectuated. Fourth, we updated both the statistical analysis plan and the methodology of the qualitative work regarding the evaluation of the PRIME Parkinson care model.

#### Extension of the evaluation period

First, the evaluation period had to be prolonged by two years because the COVID-19 crisis delayed the implementation of the new care model. Specific examples include the absence of personnel due to illness, restrictions in physical meetings during the lockdown, and slowing of the development of innovations with PwP and their caregivers. We therefore assume that PRIME Parkinson care was ineffective in the first year of evaluation (2021).

The COVID-19 crisis also caused a delay in the recruitment process of the PRIME-NL questionnaire study. In total, 984 PwP (414 in innovation region; 570 in usual care region), 566 caregivers (240 in innovation region; 326 in usual care region) and 192 healthcare professionals (67 in innovation region; 125 in usual care region) were recruited. These numbers were somewhat smaller than the original target sample size of 1200 PwP, 600 caregivers and 250 healthcare professionals.

To account for the delay and the smaller recruitment numbers, we have extended the study duration by two years (2024 and 2025). For the questionnaire-based study, we added two further follow-up assessments, resulting in a total of five follow-up measures between 2021 and 2025, i.e., one at baseline and five follow-up assessments at 12, 24, 36, 48 and 60 months. We complemented this with medical claims data collected during the same time frame. The updated power calculations given this extended study duration are presented in *Supplementary Material B*.

#### Incorporation of process measures

Since 2022, we have incorporated process measures of the implementation of the new care model in the innovation region. The implementation process was evaluated by adding specific questionnaire items which assess familiarity, usage and experiences concerning each healthcare innovation. The latter is based on the Net Promotor Score (NPS), an easy way to assess user satisfaction with one question and is therefore widely used in business and healthcare worldwide (7).

Furthermore, the roles of neurologists and Parkinson’s nurses has been shifted and extended within the new care model. This process is evaluated by extracting the number of consultations and the duration thereof from hospital systems.

In addition, we are preparing qualitative studies to obtain insight in various components of the new are model, including —but not limited to—shifts in the roles of PD nurses and neurologists and the work-life balance of healthcare professionals involved in the model. The qualitative studies will entail semi-structured interviews in a subgroup of healthcare professionals in the innovation region. We will publish the methodology of the qualitative studies separately in due time.

#### Annual feedback of interim results to stakeholders

Throughout the innovation and implementation process, we have experienced the value of incorporating key stakeholders in the development. We thereof incorporated a feedback loop of process measures and interim results of the main analyses to key stakeholders in the care model, such as healthcare innovators and healthcare professionals. The main reason is to engage those stakeholders in the care model as much as possible throughout the implementation period and beyond. This strategy is highly unusual in pharmacological studies, as feedback itself can affect the outcomes of the evaluation. By contrast, however, this strategy has previously been deployed to enhance the uptake of other healthcare innovations that relied in part on behavioral change, including the PD specialized allied health network *ParkinsonNet* (8, 9). The rationale for this strategy is that this information feedback loop is part of the model itself (10), and will thus also be incorporated in potential future scaling efforts following the current evaluation period. Process measures will also be fed in real-time to healthcare innovators, to finetune innovative solutions (*Figure 2*).

**Fig 2.**
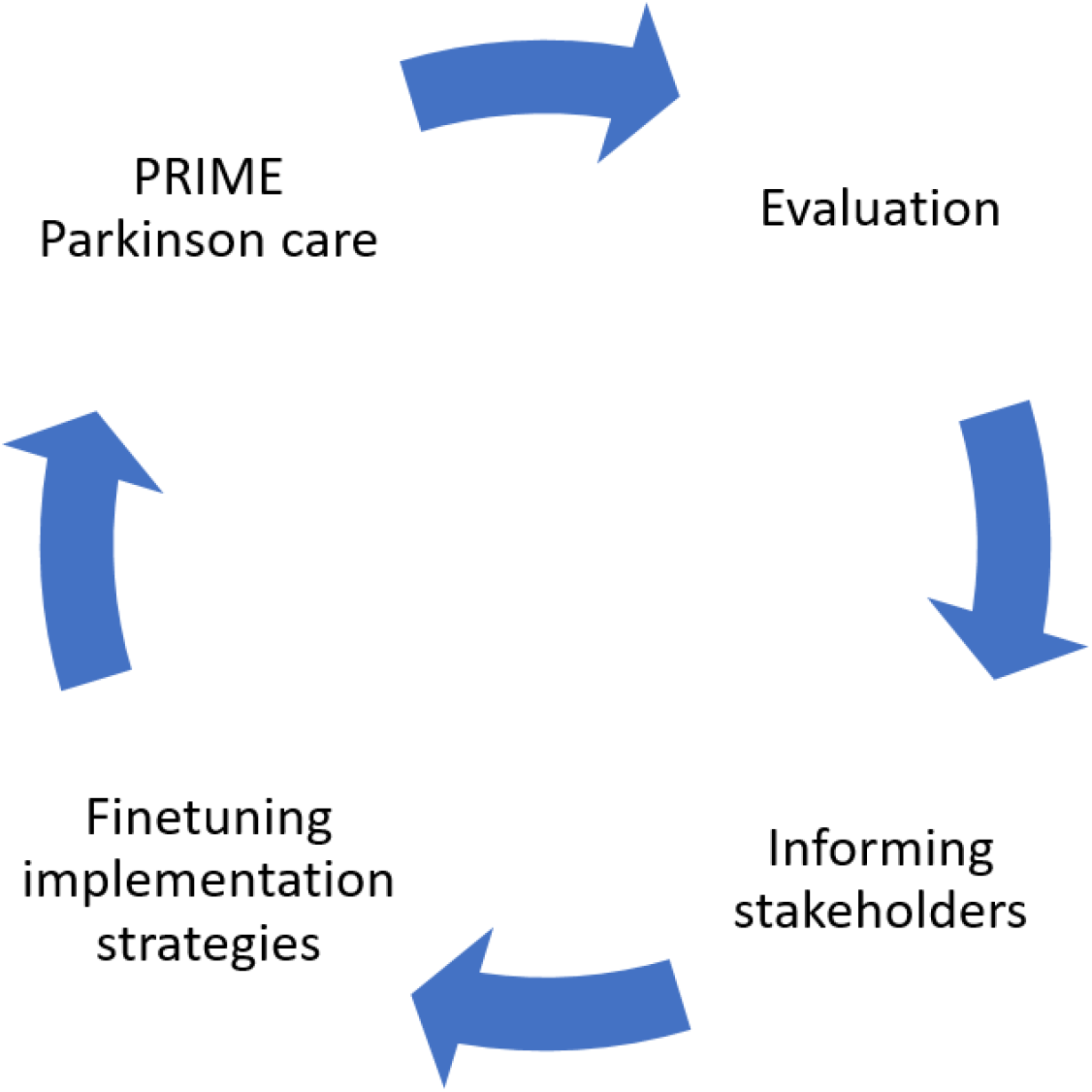
Evaluation of process indicators. From 2022, we annually evaluate process indicators, indicated by the blue-colored cycles. Based on these evaluations, we will finetune implementation strategies where necessary.

Importantly, the results of the process measure analyses and interim analyses have no bearing on the publication of the final results of the evaluation study. We will submit the results of the prespecified statistical analysis plan, which is presented in *section 2.3*, for publication no later than 1 year after completion of the final assessment year.

#### Further operationalization of outcomes

We have previously described the quantitative outcome measures both within the healthcare claims-based data and questionnaire-data of PRIME-NL study (4). The quantitative outcome measures are summarized in *Table 1*. In *Supplementary Material C and D*, we provide additional details on how the outcomes are operationalized.

**Table 1.**
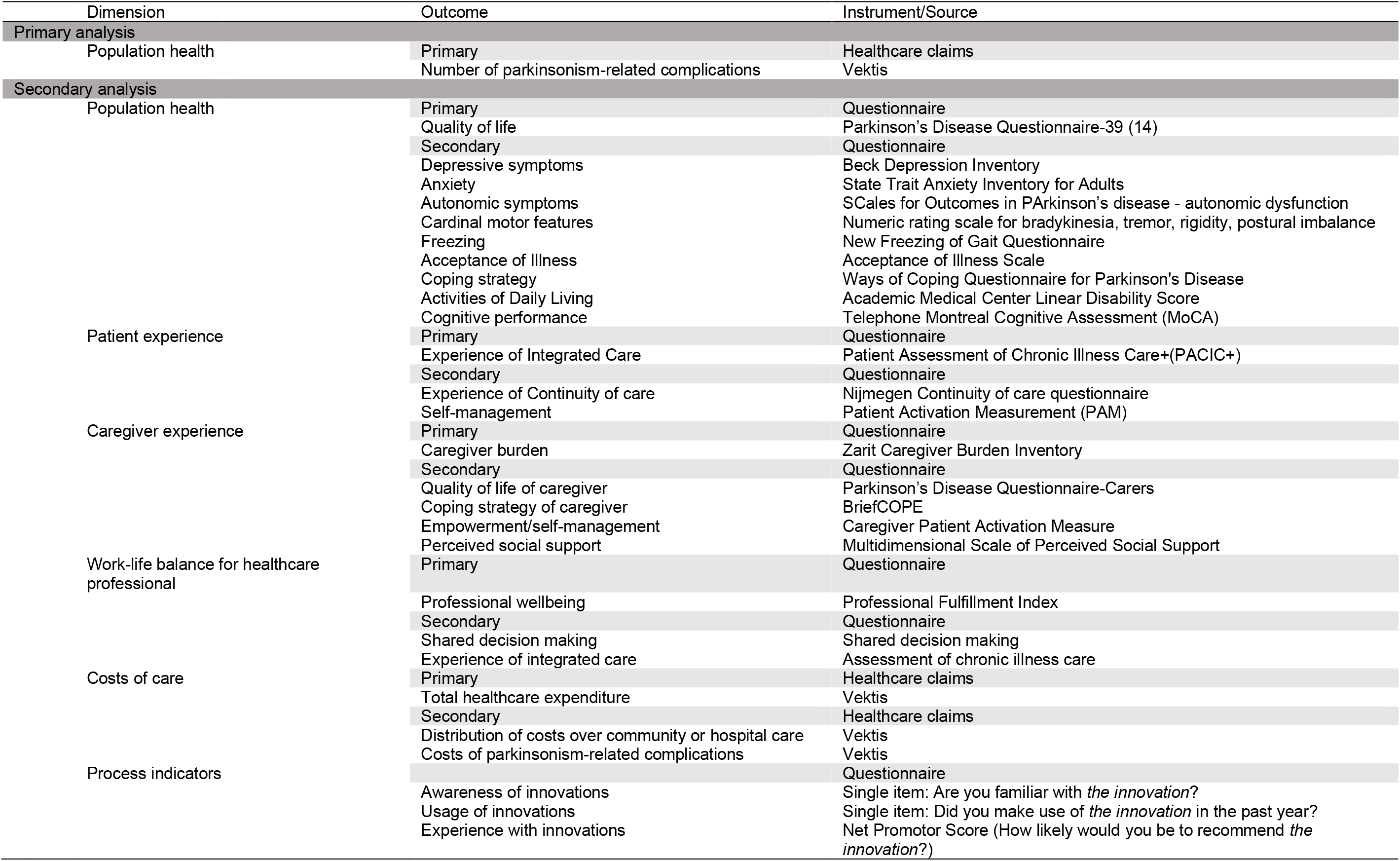
Measurable outcome measures

### 2.3 Statistical analyses

#### Primary- and secondary analyses

We previously described the statistical analyses for the primary outcome of this study, but then also indicated that we would later publish a detailed statistical analysis plan. Here, we provide this update, including our analytical approach and sample size calculations for secondary outcomes (4). Previously, we assumed the standardized difference between groups to be linear over time. However, given practice effects and delays in implementing, this assumption may not be true and either a threshold or curvilinear effect may be present.

Therefore, we will explore including exponential terms in the regression models. We previously proposed to use negative binomial regression models in the primary analysis, on the assumption that the outcome data will be over-dispersed. We will now estimate the outcome variance and if this is equal or smaller than the mean, we will use Poisson regression. Otherwise we will revert to our initial intention of negative binomial regression models. For the secondary analyses, we will use generalized mixed-effects models for all outcome measures. In the models, the region of care (innovation region or usual care region), time, and their interaction (region of care * time) are the independent variables and the outcome measure is the dependent variable. We will adjust for potential covariates (described in (4) and visualized in a DAG in (5)) as fixed effects. Random intercepts and random slopes over time for participant ID will be included. The regression coefficient and corresponding p-value for the interaction term describes the difference in annual change in the outcome measure between regions. We will consider p < .05 for the interaction term between time and region as statistically significant.

#### Sensitivity analyses

We have added several sensitivity analyses. Because of the observational nature of our study, we were not able to randomize participants between groups and therefore certain types of bias may occur. Earlier work (5) concluded that our groups slightly differ in terms of age and disease duration and therefore we will use a propensity score matching procedure in sensitivity analyses.

Also, we will repeat the main analysis after incorporating inverse probability weighting to account for regional differences in population characteristics, to quantify the influence of potential selective inclusion of participants by region as much as possible.

We are aware of changing healthcare strategies and innovations in both the innovation and control region, with possible “contamination” due to our pragmatic real world evaluation. For example, in other areas of the Netherlands, a different innovative healthcare model has been introduced which focuses on intensive inpatient rehabilitation of patients who are on the brink of permanent nursing home admission (11). Therefore, we will repeat the main analysis on the population health dimension, in which we will exclude people receiving other complex healthcare interventions that we are aware of in the Netherlands as a sensitivity check.

Separately, we will explore the intervention effect as a time-varying covariate.

For the costs of care dimension, we will exclude university medical centers, because they perform more expensive treatments are not equally distributed across regions. Another sensitivity analysis will use not the actual costs of care (as primary outcome in the costs of care dimension), but compute total costs as claims volume times average national claims prices to correct for any price differences between hospitals.

Lastly, we will perform an exploratory analysis in which we repeat the main analysis in the patient- and caregiver experience dimension, but excluding people in the innovation region who reported not using any healthcare innovations. We are aware that this is a biased approach to evaluate the utility of PRIME Parkinson care. However, this analysis serves to gain insight in the effect of the information resources on patient- and caregiver experience dimension rather than evaluating the utility of PRIME Parkinson care as a whole.

## 3. Discussion

This paper provides a transparent overview of how the design and analyses of the PRIME-NL study (4) have been adapted to control for external influences in a dynamic real world environment. For example, since the introduction of our healthcare initiative, a devastating pandemic occurred which necessitated a development of new health care strategies in both the innovation- and control regions. Another example was the introduction of complimentary healthcare innovations in other parts of the Netherlands, which serve as a control region for our innovation region – these may dilute the contrast between the innovation and control arms in our study. Typically, researchers do not make major changes to a study design after the start of the study as this is typically regarded as scientifically unsound. Making late adjustments to a study protocol while the study is already ongoing could be interpreted as data dredging (12). Being an observational study, PRIME-NL is situated within an ever-changing environment that dynamically responds to such external influences. Instead of regarding it as a weakness, we leveraged the observational nature of this study to adapt and improve the study’s design to suit its dynamic environment. With these changes, however, also comes the responsibility to make any decisions and planned analyses transparent to ensure scientific integrity and to enhance the validity of our conclusions. We have committed to reporting future updates regarding the study’s design and analyses on the Open Science Framework. This approach might serve as a template for -and could inspire-other studies on healthcare innovations.

Although we aimed to optimize the study design, there are still some remaining limitations in our updated design. The observational nature introduces for example selection bias by the inability to randomize participants (5). However, PRIME Parkinson care has also been operationalized in the Bath area in the United Kingdom (PRIME-UK) as a single-centre open label randomised controlled trial (13).This enables us to investigate the utility of PRIME Parkinson care both in a real-life situation (in the Netherlands), as well as in a controlled environment (in the United Kingdom) and triangulate our results. Another limitation might be any underestimation in the inflation of the cost of care dimension. At the moment, we assume a discount rate of 3% as advised by Dutch economic evaluation guidelines (14), in sensitivity analyses we explore discounting costs according to the Dutch medical consumer price index (15).

Taken together, the evaluation of a complex healthcare model that is implemented in the real world with external “perturbations” introduces challenges but also allowed us to adapt and improve our study design. This approach may be used in the design of a similar model for other chronic health conditions. Our solutions and transparent descriptions could both serve as a template for other complex healthcare evaluations.

## Data Availability

All data produced in the present study are available upon reasonable request to the authors

## Ethical Compliance Statement

The PRIME-NL study has been approved by the Ethical Board of the Radboud University Medical Center, reference number 2019-5618. The study has therefore been performed in accordance with the ethical standards laid down in the 1964 Declaration of Helsinki and its later amendments. Participants signed a digital or written informed consent before inclusion in the study.

We confirm that we have read the Journal’s position on issues involved in ethical publication and affirm that this work is consistent with those guidelines.

