## Supplementary Information for "The PRIME-NL study: evaluating a complex healthcare intervention for people with Parkinson’s disease in a dynamic environment"

**Supplementary material A**. Home, spoke and hub model (Adapted from (1))

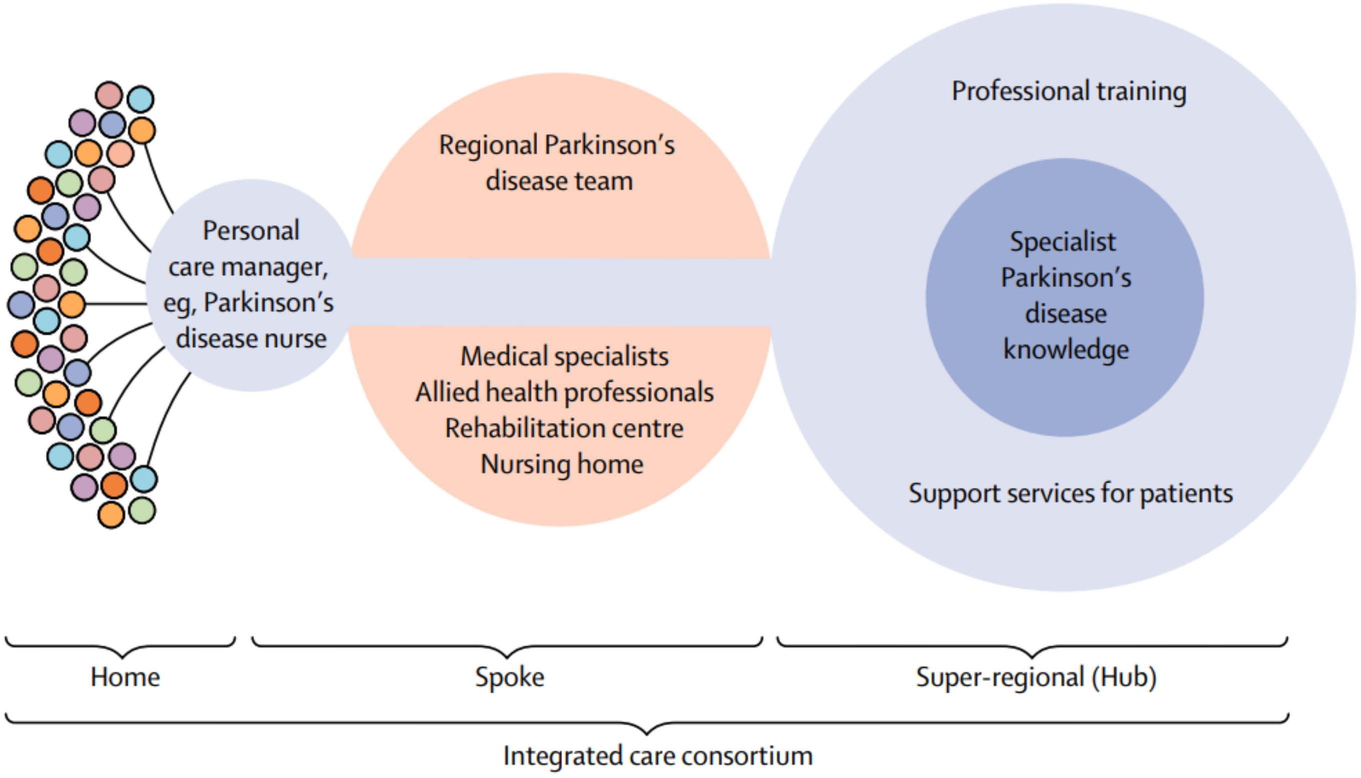


PRIME Parkinson care is operationalized using the home, spoke and hub model and comprises the following core elements: (a) specialized Parkinson's nurses who work for the patient's total care network and who play an important role in the coordination and integration of care; (b) regional teams, including specialist neurologists and a network of specially trained allied health professionals (ParkinsonNet); (c) an expertise center that supports Parkinson's nurses and regional teams; and (d) self-management by well-informed patients. This complete model is supported by various technology products and a range of centralized services to support patients and professionals.

**Supplementary material B**. Power calculations

Because of the delay of implementation of PRIME Parkinson care and the decision to extend the evaluation period by two years, we present updated power calculations. In all power calculations, we assume that PRIME Parkinson care was not effective during the first year of implementation (2021). We expect that the relative standardized difference in outcome measures after five years will be 133% (four years instead of three years) of the original expected standardized difference. We used an alpha of 5% in all power calculations. We expect an attrition of 10% of the participants in the questionnaire-based study after 3 years, and an additional 5% attrition of participants per additional extension year.

Although the exact estimation of potential effect sizes of PRIME Parkinson care are challenging due to differences in context, design, and healthcare interventions with other studies, we based our predictions on research about reasonably similar care innovations. The expected relative standardized difference in complications of 15% was based on a similar finding in a comparison between specialized physiotherapy to generic physiotherapy in a previous healthcare claims database study (2). For both the dimensions population health and patient experience, a relative 15% difference between the regions was based on a similar German study that investigated an integrative care model (3). A relative 25% standardized difference between regions in caregiver experience was based on an a study that found that home-based, individualized occupational therapy was associated with a relative 25% reduction in caregiver burden compared to usual care without occupational therapy (4).

For the updated power calculations, we assumed that the difference between the two regions in all outcome measures would be linear over time. We are aware that a ceiling or floor effect may occur, but we assume that this ceiling effect will occur after the PRIME-NL study has ended because the impact of health innovations on health outcomes takes time. PRIME Parkinson care is not disease modifying, however we hypothesize that by providing more proactive care, both the number of and severity of complications and admissions will be reduced. An endpoint in which a floor effect might occur is the number of complications, where someone cannot have less than zero complications. Furthermore, outcomes on questionnaires are bounded by a minimum and maximum score, prohibiting limitless linear change.

| Dimension | Expected difference after 3 years | Expected difference after 5 years | Power to detect a change after 5 years |
|---|---|---|---|
| Primary analysis | | | |
| Population health | 15% | 20% | 99% |
| Secondary analysis | | | |
| Population health | 15% | 20% | 87% |
| Patient experience | 15% | 20% | 87% |
| Caregiver experience | 25% | 33% | 96% |
| Healthcare professional experience | 35% | 47% | 87% |

Expected differences in outcome measures between innovation region and usual care region are presented after both 3 and 5 years of implementation. Furthermore, power of the updated power calculations is presented.

**Supplementary material A**. Definition of parkinsonism-related complications

The primary endpoint is the number of parkinsonism-related complications when comparing the innovation region to the control region. This reflects the population health dimension of the quadruple aim (1). The nationwide healthcare claims-based database from Vektis will be used to determine parkinsonism-related complications, which is defined as one of the following events: sustaining a fracture or other orthopedic injury, urinary tract infection, pneumonia or neuropsychiatric disorders, such as psychosis, depression, anxiety, or dementia. (see table below for overview of all diagnostic codes that will be used to identify parkinsonism-related complications). Each event is weighted equally. Events which fulfill these criteria are parkinsonism-related complications irrespective of whether they require hospitalization or were caused by parkinsonism. For instance, an elbow fracture (orthopedic injury) may be diagnosed at an emergency department but not lead to hospitalization. If a PwP sustains multiple parkinsonism-related complications within one month, only one complication is counted. This will also be done when two complications are present at the same timepoint. The reason for this is that we hypothesize that PRIME Parkinson care is more likely to prevent a complication that occurs in the person's home environment rather than to prevent a second or third complication that develops as a consequence of the first complication at the hospital. For instance, a PwP may fall and sustain a hip fracture in their home environment which triggers admission to the hospital, where secondary complications such as a urinary tract infection due to immobilization or a hospital delirium may occur. In this example, only the hip fracture is counted as a complication.

| Complication group | Specialty category | Vektis diagnostic code | Complication / Diagnosis |
|---|---|---|---|
| Orthopedic injuries | | | |
| | Surgery | 0303-0205 | Clavicle |
| | Surgery | 0303-0207 | Humerus proximal and shaft |
| | Surgery | 0303-0208 | Distal humerus/ (epi)condyle(s) |
| | Surgery | 0303-0210 | Radius head |
| | Surgery | 0303-0211 | Forearm nno |
| | Surgery | 0303-0212 | Wrist |
| | Surgery | 0303-0216 | Ribs, sternum |
| | Surgery | 0303-0217 | Pelvis/ sacrum |
| | Surgery | 0303-0218 | Femur, proximal (+ collum) |
| | Surgery | 0303-0219 | Femur other |
| | Orthopedics | 0305-1703 | Loosening/ infection/ malposition of pelvis/ hip/ thigh prosthesis |
| | Orthopedics | 0305-3003 | Sternum/ ribs |
| | Orthopedics | 0305-3006 | Clavicle |
| | Orthopedics | 0305-3008 | Humerus proximal and shaft |
| | Orthopedics | 0305-3009 | Distal humerus/ (epi) condyle (s) |
| | Orthopedics | 0305-3011 | Radius cup |
| | Orthopedics | 0305-3012 | Forearm |
| | Orthopedics | 0305-3013 | Wrist |
| | Orthopedics | 0305-3017 | Pelvis |
| | Orthopedics | 0305-3018 | Acetabulum |
| | Orthopedics | 0305-3019 | Femur proximal (+ collum) |
| | Orthopedics | 0305-3020 | Femur other |
| | Orthopedics | 0305-3207 | Hip, prosthesis |
| Pneumonia | | | |
| | Internal med | 0313-0401 | Pneumonia nno |
| | Lung diseases | 0322-1401 | Pneumonia |
| | Geriatrics | 0335-273 | Pneumonia |
| Urinary tract infections | | | |
| | Urology | 0306-082 | Interstitial cystitis |
| | Urology | 0306-032 | Bladder infection |
| | Internal medicine | 0313-421 | Urinary tract infection |
| | Geriatrics | 0335-301 | Disorders of the genitourinary system |

| | | | |
|---|---|---|---|
| Neuropsychiatric disorders | | | |
| | Psychiatrists | 0329-02 | Delirium, dementia and amnestic and other cognitive disorders |
| | Psychiatrists | 0329-05 | Schizophrenia and other psychotic disorders |
| | Psychiatrists | 0329-06 | Mood disorders |
| | Psychiatrists | 0329-07 | Anxiety disorders |
| | Geriatrics | 0335-241 | Mental disorders |
| | Geriatrics | 0335-242 | Memory problems and dementia |
| | Geriatrics | 0335-243 | Delirium |
| | Geriatrics | 0335-244 | Depressive disorders |
| | Geriatrics | 0335-245 | Behavioural disorders |
| | Revalidation | 0327-0716 | Mental disorders |

Diagnostic codes will be used to select complications in medical claims database Vektis

**Supplementary material D**. Outcome measure: costs of care

Endpoints within the costs of care dimension are collected from nationwide healthcare claims-data source Vektis, capturing all health contacts from primary and community care. For this dimension, we primarily focus on total healthcare expenditure. Secondary outcomes are the distribution between community care expenditure and general hospital care expenditure, and parkinsonism-specific hospital care expenditure. The latter is defined as hospital costs that are made as part of a hospital admission of which the registered reason is parkinsonism itself or one of the four above-mentioned Parkinson-specific complications, as registered in the claims database. Because costs and effect (complications) occur in different years, we will use a discount rate of 4% for costs and 1.5% for health effects (as advised by the Dutch guideline for economic evaluations) to derive net present value, similar to an earlier approach (2).
